# Advancing our understanding of skeletal muscle across the lifecourse: protocol for the MASS_Lifecourse study and characteristics of the first 80 participants

**DOI:** 10.1101/2021.10.14.21264985

**Authors:** R M Dodds, C Hurst, S J Hillman, K Davies, T J Aspray, A Granic, A A Sayer

**Affiliations:** AGE Research Group, Newcastle University Institute for Translational and Clinical Research, Newcastle upon Tyne, UK; NIHR Newcastle Biomedical Research Centre, Newcastle University and Newcastle upon Tyne NHS Foundation Trust, Newcastle upon Tyne, UK

## Abstract

**Introduction:** Sarcopenia, the age-related loss of skeletal muscle strength and mass, carries a significant burden for affected individuals. There has been little investigation of sarcopenia using experimental medicine techniques to study human muscle tissue in detail. The aim of the Muscle Ageing Sarcopenia Studies Lifecourse (MASS_Lifecourse) study is to recruit up to 160 participants, equally divided between females and males between ages 45 and 85 years for detailed phenotyping of skeletal muscle health. Here we describe the protocol for the study and the characteristics of the first 80 participants.

**Methods:** We are recruiting participants from three sources in the north-east of England. Study fieldwork comprises a home visit (or videocall) for consent and assessment of health, cognition, lifestyle, and wellbeing. This is followed by a visit to a clinical research facility for assessment of sarcopenia status and collection of samples including a vastus lateralis muscle biopsy. We produced descriptive statistics for the first 80 participants, including expressing their grip strength relative to normative data in the form of Z-scores.

**Results:** The first 80 participants (53.8% female) covered the target ages, ranging from 48 to 84 years. They were regularly physically active, reported good physical function and had a prevalence sarcopenia (including probable sarcopenia) of 11.3% based on the revised European consensus. Their grip strength was similar to that in the general population, with a mean Z-score of 0.09 standard deviations (95% CI: -1.64, 1.83) above that expected.

**Conclusions:** The MASS_Lifecourse study combines comprehensive health and lifestyle data with a range of biological samples including skeletal muscle. The findings from planned analyses should contribute to improvements in the diagnosis, treatment, and prevention of sarcopenia.

## Introduction

Sarcopenia is the loss of muscle strength and mass as we age. It refers specifically to the situation where a person’s muscle strength, from tests such as grip strength and five chair stand time, and mass, from measurements such as a dual-energy X-ray absorptiometry (DXA) scanning, are below standard cut-points [1]. The importance of muscle strength and mass is highlighted by the strong evidence that people with lower results for these tests are at greater risk of disability [2], use of social and health care including hospitalisation [3,4] and even all-cause mortality rates [5].

There is therefore growing clinical interest in sarcopenia and its potential treatments [6]. A range of mechanisms for sarcopenia have been investigated including mitochondrial dysfunction, anabolic resistance, cellular senescence, motor unit remodelling and oxidative stress [7]. However to date there has been comparatively little investigation of sarcopenia using recent experimental medicine techniques such as the use of ‘omics’ to study human muscle tissue in detail [8].

The prevalence of sarcopenia increases with age, with around one-quarter of those aged 85 found to be affected in data from the Newcastle 85+ Study [9]. By this age, it is more likely that sarcopenia will be accompanied by other ageing syndromes such as disability and frailty. This may limit our ability to establish whether changes seen in muscle are due to sarcopenia, as they may reflect both sarcopenia and its associated conditions.

An alternative approach is to explore mechanisms of skeletal muscle ageing in the period spanning both mid-life and old age, defined in this study as ages 45-85 years. This will provide opportunities to better understand risk factors and to be able to implement interventions at an earlier stage, analogous to the screening in mid-life that is now established for cardiovascular disease risk [10]. An added benefit of recruiting people in this age group for detailed assessment of skeletal muscle is that participants who may be eligible for sarcopenia trials can be quickly identified [11].

We have previously carried out two pilot Muscle Ageing Sarcopenia Studies (MASS) of detailed muscle phenotyping (MASS_Pilot [12] and MASS_PD [13]), and we have built on this experience to design the MASS_Lifecourse study. The overall aim of the MASS_Lifecourse study is to recruit a sample of up to 160 participants, divided between women and men between ages 45 and 85 years for detailed phenotyping of skeletal muscle health. In the present paper we describe (i) the protocol for the MASS_Lifecourse study and (ii) the characteristics of the first 80 participants.

## Methods

### Recruitment

We recruited participants from the north-east of England from three different sources. These were: primary care (local GP practices acting as patient identification centres within the North East and North Cumbria Clinical Research Network), secondary care (clinics run within the Newcastle upon Tyne Hospitals NHS Foundation Trust), and the NIHR Bioresource Centre Newcastle (a large panel of individuals who have expressed an interest in taking part in research studies). We started the fieldwork in October 2018, with suspension between March 2020 and September 2020 due to the COVID-19 pandemic.

#### Inclusion and exclusion criteria

The inclusion criteria were being aged between 45 and 85 years and having capacity to consent to take part. The exclusion criteria were taking anticoagulant or antiplatelet medications (except for aspirin for primary prevention of cardiovascular disease which could be suspended), diabetes mellitus, immunosuppressant medication, and pregnancy. We also excluded individuals where biopsy of the vastus lateralis muscle was judged not to be feasible by a clinician in the team. Prompts for further assessment by a clinician include a body mass index (BMI) in the obese range, significant mobility limitation or conspicuous superficial veins at the biopsy site. We assessed these criteria throughout the recruitment process to identify at an early stage if a person was not eligible to take part.

#### Ethical approval and informed consent

The study was approved by the North East – Newcastle & North Tyneside 1 Research Ethics Committee. We obtained informed consent from all participants, including asking if they gave permission for biological samples to be used in future collaboration with partners outside the UK and those in the commercial sector. Consent was rechecked before each part of the study.

### Study fieldwork

The study fieldwork comprised (1) consideration of inclusion and exclusion criteria (as above); (2) assessment of health, cognition, lifestyle and wellbeing, carried out either in the participant’s home or via videocall (with the latter introduced following the COVID-19 pandemic); followed by a visit to CARU (our Clinical Ageing Research Unit: a clinical research facility) for (3) identification of sarcopenia status and other clinical measurements and (4) collection of skeletal muscle and other samples. A summary of the fieldwork components is given in Table 1 and full details are provided below.

**Table 1:** Overview of study fieldwork.

| Component | Location | Components |
| --- | --- | --- |
| Initial contact to discuss study | Telephone | • Opportunity to raise questions after reading study participant information sheet<br>• Assessment of inclusion and exclusion criteria |
| Assessment of health, cognition, lifestyle and wellbeing | Own home* | • Long-term conditions and medications<br>• SMMSE and MoCA<br>• Food frequency questionnaire<br>• RAPA and GeneActiv 7-day wrist-worn accelerometry<br>• Smoking and alcohol<br>• Education and occupation<br>• Home ownership and number of cars<br>• Geriatric depression scale<br>• SF-36 |
| Assessment of sarcopenia status | CARU | • SARC-F<br>• Hand grip strength<br>• Short physical performance battery: chair stand test, gait speed, balance tests<br>• Segmental bioimpedance<br>• DXA |
| Other clinical measurements | CARU | • Height and weight<br>• Hip and waist circumference<br>• Lying and standing blood pressure |
| Collection of skeletal muscle and other samples | CARU | • Vastus lateralis muscle biopsy<br>• Blood samples (fasting)<br>• Urine sample<br>• Collection of stool sample kit |

\* With some conducted via videocall from September 2020 onwards.
CARU, Clinical Ageing Research Unit. DXA, dual-energy X-ray absorptiometry. MoCA, Montreal cognitive assessment. SF-36, Short Form 36 questionnaire. SMMSE, standardized mini-mental state examination. RAPA, rapid assessment of physical activity questionnaire.

#### Assessment of health, cognition, lifestyle and wellbeing

We collected information on health status, including long-term conditions, regular medications, hospital admissions and contact with health and social care professionals. We assessed cognition using the standardized mini-mental state examination (SMMSE) [14] and Montreal cognitive assessment (MoCA) [15]. We assessed smoking history, alcohol intake, and diet using a 20-item food frequency questionnaire [16]. We characterised habitual physical activity in two ways: firstly, using the rapid assessment of physical activity (RAPA) questionnaire [17] and secondly with seven-day accelerometery using a GENEActiv® Original physical activity monitor (ActivInsights Ltd, Cambridge, UK) worn on the dominant wrist at a measurement frequency of 100 Hz.

We asked participants about their educational history (years spent in school and in higher education), home and car ownership, and their own and (if relevant) their partner’s main occupation classified using the Office for National Statistics (ONS) simplified NS-SEC analytic class [18]. We assessed quality of life using the Short Form 36 (SF-36) questionnaire [19] and mood using the Geriatric Depression Score [20].

#### Assessment of sarcopenia status and other clinical measurements

We assessed sarcopenia status using measures recommended in the revised European consensus on the definition and diagnosis of sarcopenia [1]. We assessed self-reported physical function using the SARC-F questionnaire [21]. We measured grip strength (kg) by taking three assessments from each hand using a Jamar dynamometer, with the maximum value used in analyses [22]. We recorded the time take to complete five chair stands [23]. We tested physical performance by measuring normal gait speed over 4 metres and tests of standing balance. We assessed appendicular lean mass using segmental bioimpedance using a Tanita MC-780MA body composition analyser (Tanita Corporation, Arlington Heights, IL, USA), and using DXA (Lunar iDXA, GE Healthcare, USA). Other clinical measurements included height, weight, hip and waist circumference, and lying and standing blood pressure.

#### Collection of skeletal muscle and other samples

Participants collected a stool sample at home using the OMNIgene GUT kit (DNA Genotek, Ottawa, Canada) and brought this with them. Once in CARU we collected blood samples after an overnight fast, used to perform a set of routine clinical tests, as well as to provide stored blood for subsequent DNA and RNA extraction and aliquots of serum and plasma.

After participants had breakfast, we asked them to provide a urine sample for subsequent metabolomic analyses. At approximately 11am on the day of the CARU visit we performed a biopsy of the vastus lateralis muscle, usually taken on the right side, and using a Weil-Blakesley conchotome [24]. We aimed to collect three muscle samples, with one placed immediately into RNAlater solution (RNAlater™ stabilization solution, Thermofisher Scientific). The other two were placed in gauze prior to processing: one sample orientated and mounted for histology, and then both samples flash frozen in isopentane cooled in liquid nitrogen prior to storage at -80°C [25]. We observed the participant for around two hours following the biopsy procedure and followed-up by telephone the next day and a week later.

### Data management

We stored participants’ details in a password-protected database on a secure network drive, accessible only by members of the study team with the required approvals in place. We used a pseudonymised participant identifier throughout the fieldwork. We used double data entry when inputting data from paper forms. We carried out checks for values outside of the feasible range, and we checked the consistency when information was combined from different sources (for example, checking that a participant’s weight recorded during a DXA scan was consistent with that from bioimpedance analysis).

### Maintenance of cohort

At the end of the fieldwork, we sought participants’ consent to keep them updated on progress with the study and we send an annual newsletter for this purpose. We also sought their permission to contact them about further studies that might be of interest; to date, this has included attending for magnetic resonance imaging of skeletal muscle to assess motor unit function [26].

### Statistical analyses in this paper

In the present paper, we describe the characteristics of the first 80 participants, based on a selection of the variables described under study fieldwork, above, and as shown in Table 2.

**Table 2:**
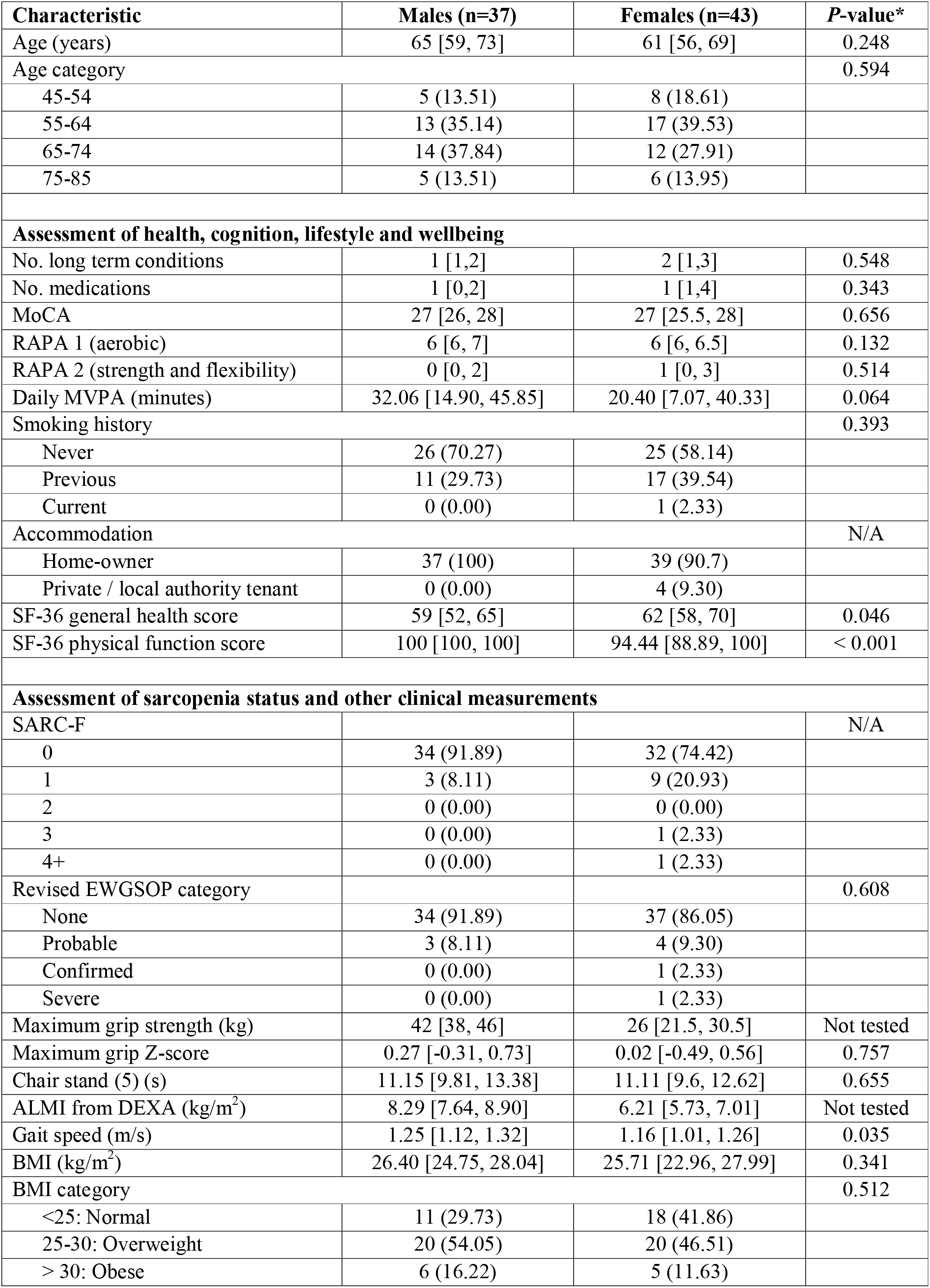
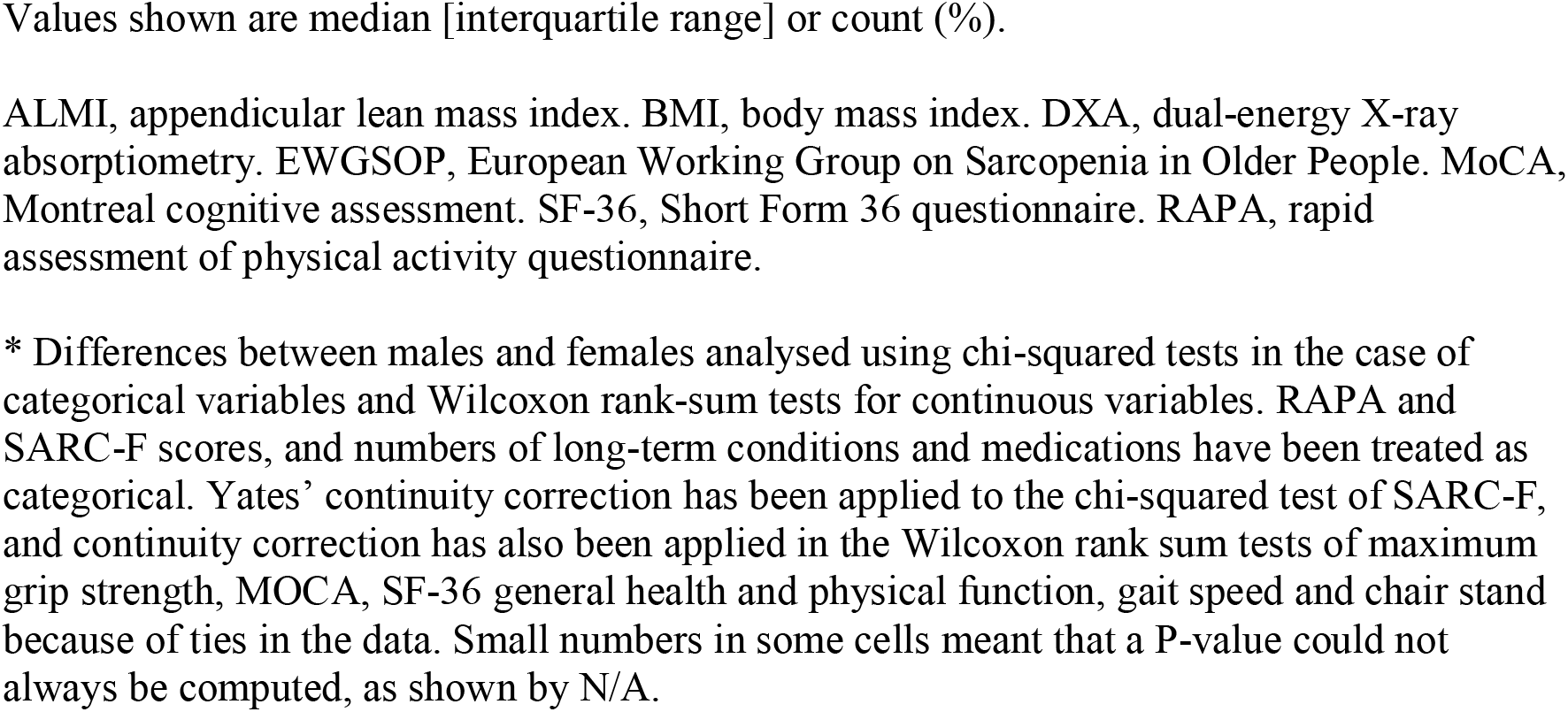
Characteristics of the sample, by sex.

| Characteristic | Males (n=37) | Females (n=43) | P-value* |
| --- | --- | --- | --- |
| Age (years) | 65 [59, 73] | 61 [56, 69] | 0.248 |
| Age category |  |  | 0.594 |
| 45-54 | 5 (13.51) | 8 (18.61) |  |
| 55-64 | 13 (35.14) | 17 (39.53) |  |
| 65-74 | 14 (37.84) | 12 (27.91) |  |
| 75-85 | 5 (13.51) | 6 (13.95) |  |
| **Assessment of health, cognition, lifestyle and wellbeing** |  |  |  |
| No. long term conditions | 1 [1,2] | 2 [1,3] | 0.548 |
| No. medications | 1 [0,2] | 1 [1,4] | 0.343 |
| MoCA | 27 [26, 28] | 27 [25.5, 28] | 0.656 |
| RAPA 1 (aerobic) | 6 [6, 7] | 6 [6, 6.5] | 0.132 |
| RAPA 2 (strength and flexibility) | 0 [0, 2] | 1 [0, 3] | 0.514 |
| Daily MVPA (minutes) | 32.06 [14.90, 45.85] | 20.40 [7.07, 40.33] | 0.064 |
| Smoking history |  |  | 0.393 |
| Never | 26 (70.27) | 25 (58.14) |  |
| Previous | 11 (29.73) | 17 (39.54) |  |
| Current | 0 (0.00) | 1 (2.33) |  |
| Accommodation |  |  | N/A |
| Home-owner | 37 (100) | 39 (90.7) |  |
| Private / local authority tenant | 0 (0.00) | 4 (9.30) |  |
| SF-36 general health score | 59 [52, 65] | 62 [58, 70] | 0.046 |
| SF-36 physical function score | 100 [100, 100] | 94.44 [88.89, 100] | < 0.001 |
| **Assessment of sarcopenia status and other clinical measurements** |  |  |  |
| SARC-F |  |  | N/A |
| 0 | 34 (91.89) | 32 (74.42) |  |
| 1 | 3 (8.11) | 9 (20.93) |  |
| 2 | 0 (0.00) | 0 (0.00) |  |
| 3 | 0 (0.00) | 1 (2.33) |  |
| 4+ | 0 (0.00) | 1 (2.33) |  |
| Revised EWGSOP category |  |  | 0.608 |
| None | 34 (91.89) | 37 (86.05) |  |
| Probable | 3 (8.11) | 4 (9.30) |  |
| Confirmed | 0 (0.00) | 1 (2.33) |  |
| Severe | 0 (0.00) | 1 (2.33) |  |
| Maximum grip strength (kg) | 42 [38, 46] | 26 [21.5, 30.5] | Not tested |
| Maximum grip Z-score | 0.27 [-0.31, 0.73] | 0.02 [-0.49, 0.56] | 0.757 |
| Chair stand (5) (s) | 11.15 [9.81, 13.38] | 11.11 [9.6, 12.62] | 0.655 |
| ALMI from DEXA (kg/m<sup>2</sup>) | 8.29 [7.64, 8.90] | 6.21 [5.73, 7.01] | Not tested |
| Gait speed (m/s) | 1.25 [1.12, 1.32] | 1.16 [1.01, 1.26] | 0.035 |
| BMI (kg/m<sup>2</sup>) | 26.40 [24.75, 28.04] | 25.71 [22.96, 27.99] | 0.341 |
| BMI category |  |  | 0.512 |
| <25: Normal | 11 (29.73) | 18 (41.86) |  |
| 25-30: Overweight | 20 (54.05) | 20 (46.51) |  |
| > 30: Obese | 6 (16.22) | 5 (11.63) |  |

Values shown are median [interquartile range] or count (%).
ALMI, appendicular lean mass index. BMI, body mass index. DXA, dual-energy X-ray absorptiometry. EWGSOP, European Working Group on Sarcopenia in Older People. MoCA, Montreal cognitive assessment. SF-36, Short Form 36 questionnaire. RAPA, rapid assessment of physical activity questionnaire.
\* Differences between males and females analysed using chi-squared tests in the case of categorical variables and Wilcoxon rank-sum tests for continuous variables. RAPA and SARC-F scores, and numbers of long-term conditions and medications have been treated as categorical. Yates' continuity correction has been applied to the chi-squared test of SARC-F, and continuity correction has also been applied in the Wilcoxon rank sum tests of maximum grip strength, MOCA, SF-36 general health and physical function, gait speed and chair stand because of ties in the data. Small numbers in some cells meant that a P-value could not always be computed, as shown by N/A.

From the GeneActiv physical activity monitor recordings we calculated the mean daily duration of moderate to vigorous physical activity (MVPA) using the library GGIR [27], defined as periods for which measured acceleration was greater than 0.1*g* calculated using the Euclidean norm minus one algorithm, and sustained for at least 80% of a minimum bout of 5 minutes.

We used the cut-points recommended in the revised European consensus guidelines for diagnosis and severity of sarcopenia [1], classifying participants as having probable sarcopenia (weak grip strength [< 27 kg in males and < 16 kg in females] and/or prolonged time for five chair stands [> 15 seconds]), confirmed sarcopenia if they additionally had low values for appendicular lean mass index (ALMI, appendicular lean mass in kg divided by height-squared [< 7.0 kg/m^2^ in males and < 5.5 kg/m^2^ in females]), and severe sarcopenia if they also had slow gait speed (≤ 0.8 m/s). We also compared our grip strength values to published British normative data [28], by expressing each patient’s value as a Z-score. Each Z-score is calculated as the patient’s grip strength value less the mean expected for their age and sex, divided by the grip strength standard deviation (SD) for their age and sex. Z-scores of +1 and -1 indicate grip strength values one standard deviation above and below, respectively, that expected for age and sex.

We produced descriptive statistics stratified by sex, with differences between males and females analysed using chi-squared tests in the case of categorical variables, and Wilcoxon rank sum tests for continuous variables. We carried out statistical analysis using R, version 4.0.4 [29].

## Results

Here we describe characteristics of the first 80 participants in the MASS_Lifecourse Study who had a home visit and attended the CARU, as shown in Table 2. The participants spanned the intended age range (minimum 48, maximum 84 and median 64 years), with an approximately even split between males (46.2%) and females (53.8%). During this period of fieldwork, we also visited seven potential participants at home and found that they had exclusion criteria that prevented them from taking part. A further five participants had a home visit and were recruited to the study but did not progress to the CARU visit due to a change in their personal circumstances.

### Assessment of health, cognition, lifestyle and wellbeing

The majority (n=45, 56.3%) of participants had two or more long-term condition (i.e. they had multimorbidity), with a tendency for females to have more long-term conditions than males, as shown in Table 2. Participants had normal cognition based on the MoCA test. In terms of lifestyle, most participants undertook regular aerobic activity based on the RAPA1 questionnaire, and in keeping with this had evidence of regular moderate-vigorous physical activity based on GENEActiv accelerometry. Most participants did not undertake regular strength and flexibility training as assessed using the RAPA2 questionnaire. Finally, most participants had high self-rated physical function based on the SF-36 questionnaire, especially so in males.

### Assessment of sarcopenia status and other clinical measurements

Most participants did not score any points on the SARC-F questionnaire (n=66, 82.5%) or meet the criteria for the European revised sarcopenia definition, with (n=9, 11.2%) having probable sarcopenia or greater, as shown in Table 2. As expected, we saw stronger average grip strength and ALMI in males compared to females. Participants’ grip strength Z-score values were typically as expected for their age and sex, with a mean Z-score of 0.09 standard deviations (95% CI: -1.64, 1.83) above population reference values. Finally, most participants had a BMI in the overweight range.

## Discussion

### Summary of findings

We have successfully established a cohort study that will improve our understanding of changes in skeletal muscle in mid-life and old age, with fieldwork ongoing at the time of writing. The assessments as described in this protocol paper include a comprehensive assessment of a range of health and lifestyle factors, cognition, and sarcopenia status. There is also collection of a range of biological samples including skeletal muscle ready for subsequent analyses including multi-omics. The first 80 participants covered the intended age range (45 to 85 years) and were characterised by having regular physical activity, good self-reported physical function, and low levels of sarcopenia.

### Interpretation of findings

We had a high rate of those who underwent a home visit going on to have assessments in the CARU (87%) as we used initial telephone contact to check exclusion criteria and to check that it was a feasible time to take part in the study. We anticipated a healthy volunteer effect and there was some evidence that the first 80 participants in MASS_Lifecourse were, on average, more active and had better self-reported physical function that the general population. In contrast, we saw levels of muscle strength (assessed by grip strength) which were at the same level as that seen in the general population, and few participants reported undertaking regular strength training.

### Strengths and limitations

A strength of our work is that we used the same recruitment sources to cover the age range from 45 – 85 years, meaning that any age-related differences in muscle characteristics will not have arisen owing to the use of different recruitment strategies in different age groups.

There was some evidence of a healthy volunteer effect, and this may have contributed to the low levels of confirmed (or severe) sarcopenia present in the study. This also has the potential to be a strength of this cohort, since it provides an opportunity to gain insights into the biological processes that lead to sarcopenia, and also to understand age-related changes in skeletal muscle (sometimes termed primary sarcopenia) separated as far as possible from muscle changes due to long-term conditions, termed secondary sarcopenia, and an area of growing research interest [1,31].

### Plans for future research

A range of analyses are planned, due to take place following the completion of study fieldwork in 2022 to avoid batch effects. This includes histology, transcriptomics, and proteomics of skeletal muscle. These will complement existing studies in this area but with a larger sample size than available previously [32]. This study protocol also forms a template for future cohort studies, including in people living with specific conditions, or clusters of long-term conditions. The use of such a standardised approach will allow the comparison of findings with data from the present study.

### Conclusions

We have successfully established a cohort study to improve our understanding of changes in skeletal muscle in mid-life and old age. The planned analyses of the samples collected in the MASS_Lifecourse study and their relation to a range of participant characteristics will considerably increase our understanding of the biological mechanisms underpinning age-related muscle loss. This knowledge should in turn lead to improvements in our ability to diagnose, treat and prevent sarcopenia.

## Data Availability

All data produced in the present study are available upon reasonable request to the authors

